# A residual marker of cognitive reserve is associated with resting-state intrinsic functional connectivity along the Alzheimer’s disease continuum

**DOI:** 10.1101/2022.01.17.22269026

**Authors:** Ersin Ersoezlue, Robert Perneczky, Maia Tatò, Julia Utecht, Carolin Kurz, Jan Häckert, Selim Guersel, Lena Burow, Gabriele Koller, Sophia Stöcklein, Daniel Keeser, Boris Papazov, Marie Totzke, Tommaso Ballarini, Frederic Brosseron, Katharina Buerger, Peter Dechent, Laura Dobisch, Michael Ewers, Klaus Fliessbach, Wenzel Glanz, John Dylan Haynes, Michael T Heneka, Daniel Janowitz, Ingo Kilimann, Luca Kleineidam, Christoph Laske, Franziska Maier, Matthias H Munk, Oliver Peters, Josef Priller, Alfredo Ramirez, Sandra Röske, Nina Roy, Klaus Scheffler, Anja Schneider, Björn H Schott, Annika Spottke, Eike Jakob Spruth, Stefan Teipel, Chantal Unterfeld, Michael Wagner, Xiao Wang, Jens Wiltfang, Steffen Wolfsgruber, Renat Yakupov, Emrah Düzel, Frank Jessen, Boris-Stephan Rauchmann, DELCODE study group

**Author notes:** Corresponding author: Dr. Boris-Stephan Rauchmann and Prof. Dr. Robert Perneczky, Division of Mental Health of Older Adults, Department of Psychiatry and Psychotherapy, Ludwig-Maximilians-Universität München, Nussbaumstr. 7, 80336 Munich, Germany. These two authors contributed equally (Ersin Ersoezlue, Robert Perneczky).

## Abstract

**Background:** Cognitive reserve (CR) explains interindividual differences in the impact of neurodegenerative burden on cognitive and daily functioning. A residual model was proposed to estimate CR more accurately compared to static measures, such as years of education. However, the functional brain correlates of residual CR markers (CRM) remain unexplored.

**Methods:** From the DELCODE cohort, 318 participants with resting-state functional and structural MRI data were included and stratified using cerebrospinal fluid (CSF) biomarkers according to the A(myloid-β, Aβ)/T(au)/N(eurodegeneration) classification scheme, resulting in 112 Aβ-negative healthy controls and 206 Aβ-positive patients in the Alzheimer’s disease (AD) spectrum.. CRM was calculated utilizing residuals obtained from a multilinear regression model using global cognition as dependent variable and demographic and disease burden measures as predictors. Associations between the CRM and intrinsic network connectivity (INC) in resting-state networks associated with cognition were explored, including the default mode network (DMN), frontoparietal network (FPN), salience network (SAL) and dorsal attention network (DAN). Moreover, the association between memory performance-associated regional INC and CRM was assessed.

**Results:** CRM was positively associated with INC in the DMN in the entire cohort. In a subgroup analysis, the A+T+N+ group revealed an anti-correlation between SAL and DMN. Furthermore, CRM was positively associated with the anti-correlation between the memory-related regions in the FPN and the DMN in the A+ and A+T/N+ subgroups.

**Conclusions:** CRM is associated with alterations of functional connectivity in resting-state networks of cognitive function, particularly the DMN and the FPN. Our results provide evidence on individual functional network differences associated with CRM in the AD continuum.

## Introduction

The concept of cognitive reserve (CR) refers to the capacity and flexibility of cognitive and brain processes that help to attenuate the impact of brain aging or pathology on cognitive function or daily activities, for example in Alzheimer’s disease (AD) (1). The related concepts of brain reserve (2) and brain maintenance describe different, complementary aspects of resilience (3). Key mechanisms underlying CR include the brain’s ability to maintain neural functions more succesfully, to recruit compensatory networks, or to use existing networks more efficiently (4).

Years of formal education (1,5) and occupational complexity (6) are used frequently as CR proxy measures (7), but they are static and only reflect selected aspects of intellectual attainment. Dynamic predictors, including residualized cognitive performance (i.e., residual CR marker (CRM)) that quantifies the discrepancy between the observed cognitive performance and the performance estimated based on the neuropathological burden of a person may offer more granular data resulting in more detailed insights into the nature of CR (3). Residual approaches were shown to be relatively reliable and were studied not only in cross-sectional (8,9) but also in longitudinal studies (10–12). A residual CR measure considers demographical and disease-related confounders multidimensionally and may therefore be more comprehensive and informative at the individual level compared to traditional markers such as education (3).

Resting-state networks (RSNs) such as the default mode network (DMN), involved in cognition, self-reference, social cognition, or autobiographical memory (i.e., inwardly directed cognition) (13–15), and networks associated with externally directed cognitive processing, including the dorsal attention (DAN), salience (SAL), and frontoparietal network (FPN) (13,16–19), are affected by AD pathology and correlate with disease progression.

Spatial links exist between AD pathology and functional connectivity (FC) changes, particularly in the posterior DMN and FPN (20). Moreover, the inter-network connectivity among RSNs is also affected in AD, especially between DMN and DAN (13) as well as SAL (21). As AD progresses, functional network changes affect predominantly intra-network connectivity, and to a lesser extent inter-network connectivity (16).

Studies showed positive associations between residual markers of CR and the graph-theoretical measurement of network efficiency and FC (8,9). FC alterations were also related to CR using a quantified proxy score considering educational attainment, mainly in DMN regions (22), and were associated with lower metabolic activity in the DMN and the DAN (23). Furthermore, increasing evidence suggests a crucial role of the FPN in CR, including global connectivity of the left frontal cortex (LPC) in resting-state fMRI, a hub region within the FPN (24–26). Compared to node-to-node connectivity analyses, whole network intrinsic connectivity analysis following the data-driven independent component analysis (27) allows FC analysis more broadly within and across networks (16).

A biologically-based definition of the AD diagnosis has been proposed in a recent research framework using a binary biomarker status (presented or absent) for (A)myloid-β (Aβ), (T)au and (N)eurodegeneration (i.e., the ATN classification) as biomarker-based diagnostic profiles, prompting the switch from a symptom-based to a biological definition of AD (28).FC alterations appear already in the preclinical and early clinical stages of AD (29), also showing meaningful effects of CR on the individual clinical progression trajectories (30,31). While the characterization of a residual CRM is improving in dementia populations (32), there is a need to operationalize them in early disease and to explore their associations with intrinsic network connectivity (INC) of cognitive RSNs. Here we aimed to examine the associations of a residual CRM with FC alterations within and between network connectivity in the AD continuum, using a biomarker-based approach for diagnosis and staging.

## Materials and Methods

Data from the prospective, observational German Center for Neurodegenerative Diseases (Deutsches Zentrum für Neurodegenerative Erkrankungen, DZNE)-Longitudinal Cognitive Impairment and Dementia Study (DELCODE) (33) was used for the present analyses.

### Participants

All eligible DELCODE participants were included if they had available clinical dementia rating (CDR), neuropsychological tests, cerebrospinal fluid (CSF) biomarker analyses and apolipoprotein E (*APOE*) genotyping results and relevant structural and functional MRI data. We classified each participant following the A/T/N classification scheme using binarized CSF biomarker measurements of Aβ for A, total tau (tTau) for N and phosphorylated-tau181 (pTau) for T (34). To restrict the cohort to participants in the AD continuum and healthy controls, we excluded participants classified as A-T/N+ (i.e., suspected non-AD pathology) and A-T-N-with a global CDR rating of higher than 0 (i.e. non-AD cognitive impairment). The final cohort of 318 participants included 112 A-T-N- individuals with global CDR=0 as healthy controls (HC, mean age 69 ± 6, 52 females) and 206 A+ patients along the AD continuum (mean age 72 ± 6, 101 females), encomassing 106 A+T-N-, 28 A+T/N+ (A+T+N- and A+T-N+) and 72 A+T+N+ individuals. The detailed inclusion and exclusion criteria and study procedures of the DELCODE study are reported elsewhere (33). The following CSF biomarker cut-off values were obtained by Gaussian mixture modeling using the R package flexmix (version 2.3-15) in the DELCODE dataset from n=527 participants (sampling rate among entire baseline cohort: 53%): Aβ42: <= 638.7 pg/ml, tTau: > 510.9 pg/ml and pTau181: >= 73.65 pg/ml.

### MR image acquisition and preprocessing

Imaging was performed at nine different DZNE sites on 3T MRI scanners (Siemens Healthineers, Erlangen, Germany; three Verio, three TimTrio, one Prisma and two Skyra) using synchronized acquisition parameters. T1-weighted anatomical imaging was acquired in a 5-minute magnetization-prepared rapid gradient echo (MPRAGE) scan with the following parameters: field of view (FOV) 256×256 □mm, isotropic voxel size: 1□mm, echo time (TE) 4.37□ ms, flip angle (FA) 7°, repetition time (TR) 2500 □ms, number of slices 192. Resting-state functional MRI was acquired in a 7-minute 54-second run (180 volumes, FOV: 224×224×165mm, isotropic voxel size: 3.5mm, TE: 30ms, TR: 2580ms, FA: 80, parallel imaging acceleration factor 2). All scans were visually inspected for completeness, cuts, subject motion and other artifacts (such as blurring, echoes and ghosting). Images were classified as usable, questionable or unusable, and only images that were classified as usable were included.

All T1-weighted images were processed in FreeSurfer (v6, http://surfer.nmr.mgh.harvard.edu/) using the recon-all pipeline, including registration to Montreal Neurological Institute (MNI) standard space, intensity normalization, brain extraction, tissue type classification, surface reconstruction and probabilistic anatomical labeling (35). Segmentations were visually checked for accuracy and corrected as needed. Cortical thickness was estimated in FreeSurfer (Desikan-Killiany) atlas segmentations. A mean cortical thickness score in a composite region comprising the most vulnerable regions to atrophy in AD was calculated. This composite score was used to adjust the subsequent analyses for inter-individual differences in the degree of cortical atrophy. The composite region included the entorhinal cortex, temporal pole, inferior and middle temporal gyri, inferior and superior parietal cortices, precuneus and posterior cingulate cortex (36).

Functional connectivity analysis was performed using the CONN-fMRI Functional Connectivity Toolbox (v17, www.nitrc.org/projects/conn) and SPM12 (www.fil.ion.ucl.ac.uk/spm/), implemented in MATLAB (Release2017b, https://de.mathworks.com/products/new_products/release2017b.html). The default preprocessing pipeline for volume-based analyses was used, comprising realignment, slice-time correction, segmentation and structural and functional normalization. The Artifact Detection Toolbox (ART)-based outlier detection (https://web.mit.edu/swg/software.htm) and smoothing using a Gaussian kernel of 6 mm at FWHM (23) was applied. Denoising was performed using the default pipeline based on linear regression of potential confounding effects of white matter and CSF (37), estimated subject-motion parameters (38), outlier scans and scrubbing, followed by applying a band-pass filter (below 0.008 Hz or above 0.09 Hz) (39). Afterward, the distribution of FC correlation values was directly compared to an associated null-hypothesis distribution that has shown a 95.5% match with null-hypothesis, indicating a lack of noticeable associations between quality control and FC (40).

### Clinical characteristics, cognitive testing and assessment of CSF biomarkers

The clinical severity of dementia symptoms was quantified using the CDR-sum of boxes (CDR-sb). Cognitive performance was assessed using the Mini-mental-state examination (MMSE) and domain-specific cognitive composite scores for episodic memory, executive functioning, visual memory, language and working memory (41). A global cognitive composite score was calculated by averaging each domain-specific score, as reported in detail elsewhere (33). CSF biomarkers were assessed using established commercially available analysis kits: V-PLEX Aβ Peptide Panel 1 (6E10) Kit (K15200E), V-PLEX Human tTau Kit (K151LAE) (Meso Scale Diagnostics LLC, Rockville, MD, USA) and Innotest Phospho-Tau(181P) (Fujirebio Germany GmbH, Hannover, Germany) (33).

### Assessment of static parameters of cognitive reserve

Individual lifestyle differences defined as static CR proxies were assessed using the total years of formal education and a validated German version (42) of the Lifetime Experiences Questionnaire (LEQ) total score, reflecting activities across the lifespan (educational, occupational, managerial history, social and intellectual activities) (43). The LEQ total score was derived as a weighted sum score of three sub-scores for different stages of life (early adulthood (LEQ-e, age 13 to 30 years), mid-life (LEQ-m, age 30 to 65 years) and late-life (LEQ-l, age 65 and older). Participants younger than 65 years were excluded from analyses comprising the LEQ total (n=132) or LEQ-l scores (n=118).

### Quantitative residual cognitive reserve marker

In order to estimate a residual CRM for each participant, we calculated a stepwise regression model including the global cognitive composite score as the dependent variable and demographic (age and sex), genetic risk and neurodegenerative burden as predictors (8), adjusting for study sites. Estimates of neurodegenerative burden included binarized *APOE* ε4 allele carrier status, CSF biomarker levels (Aβ42, tTau and pTau181), mean cortical thickness of predefined brain regions vulnerable to atrophy in AD and mean bilateral hippocampal volume (Fig. S1-A, adjusted R^2^=0.54). Sex and pTau181 level were excluded due to insignificant predictive value and high collinearity in the stepwise method.

### Independent component analysis of functional MRI

We applied an independent component analysis (ICA) to determine the spatial extent of the RSNs (27) on preprocessed resting-state fMRI data using the CONN toolbox. The number of independent components to extract was set a priori to 20 (44). To identify RSNs from the ICA components, the obtained group ICA components were spatially compared to templates derived from Yeo 7-networks (45). Following group ICA, subject-specific independent component maps of the DMN, SAL, DAN and FPN were back-reconstructed using the GICA3 algorithm (46). Subsequently, we binarized the group component maps of the according networks at a threshold of z-score >2.

### Intrinsic network functional connectivity analysis of resting-state networks

Intrinsic connectivity measures were analyzed using node centrality at each voxel, described as the strength of connectivity between a given voxel (node) and each voxel in the remainder of the grey matter. The arithmetic details of this approach, provided in the CONN-fMRI Functional Connectivity Toolbox (v17, www.nitrc.org/projects/conn), are reported elsewhere (47). The results were defined in anatomical and network regions using Harvard-Oxford Atlas (48) and binarized masks from the independent component analysis respectively.

### Seed-to-voxel functional connectivity of memory-related seed regions

Using the associations between FC and MEM (see below for statistical description), we identified regions-of-interest using binary masking based on the significant regions with MEM-related FC changes (separately) for each RSN. Masked regions were used as seed regions for every voxel in the brain. Seed-based connectivity analyses were computed using the Fisher-z-transformed bivariate correlation coefficients between a seed region’s blood-oxygen-level-dependent (BOLD) time-series and each individual voxel BOLD time-series.

### Statistical analyses

All statistical analyses were performed using SPSS, version 25.0 (IBM Corp., Somers, NY). The Bonferroni method was used to correct for multiple comparisons for analyses of demographical and clinical data, and a false discovery rate (FDR) (49) correction was applied to FC data. Kruskal-Wallis tests and Chi-square tests were used to compare baseline sociodemographic, clinical and genetic variables between the study groups. Analysis of Covariance (ANCOVA) was used to compare cortical thickness composite scores and hippocampal volumes, INC of each cognitive RSN, CSF biomarkers and CRM between the groups, adjusting for age and sex (additional adjustments were made for years of education in comparisons of cognitive assessments and for imaging sites in comparisons of INC), as appropriate.

Multilinear regression models were performed to explore the association of CRM with static CR Proxies (i.e., years of education and LEQ-total), in the whole sample and the A+ group separately, adjusted for age, sex, study sites and A/T/N diagnostic subgroups. Results were reported with standardized beta coefficients (b) considered significant when p<0.05 (two-tailed).

The associations between FC and MEM were tested, adjusting for age, sex, cortical thickness and imaging sites. Similarly, the associations between CRM and INC in the resting-state FC networks were tested at voxel-level using general linear models (separately) in the entire sample, in A+, and in A/T/N subgroups. Likewise, the associations between CRM and any MEM-related connectivity seeds for each resting-state network were (separately) tested using general linear models (separately) in the A+ and in A/T/N groups. Models were adjusted for age, sex, site, cortical thickness composite score and in the A+ also for A/T/N group. All models were adjusted for age, sex, imaging site, cortical thickness and A/T/N status. Results were considered significant when p<0.05 in Gaussian random field theory (50) for INC, indicating a significance when cluster-level FDR-corrected p<0.05 and voxel-level p<0.001.

## Results

The characteristics of the study groups are shown in **Table 1**. HC were younger than A+ participants, while the groups did not differ in years of education or CRM. A+ individuals were more frequently *APOE* ε4 allele carriers, had lower mean hippocampal volumes and mean cortical thickness, lower CSF Aβ42, higher CSF tTau, and pTau181, higher global CDR scores and lower MMSE as well as lower global cognitive composite scores, as expected.

**Table 1.**
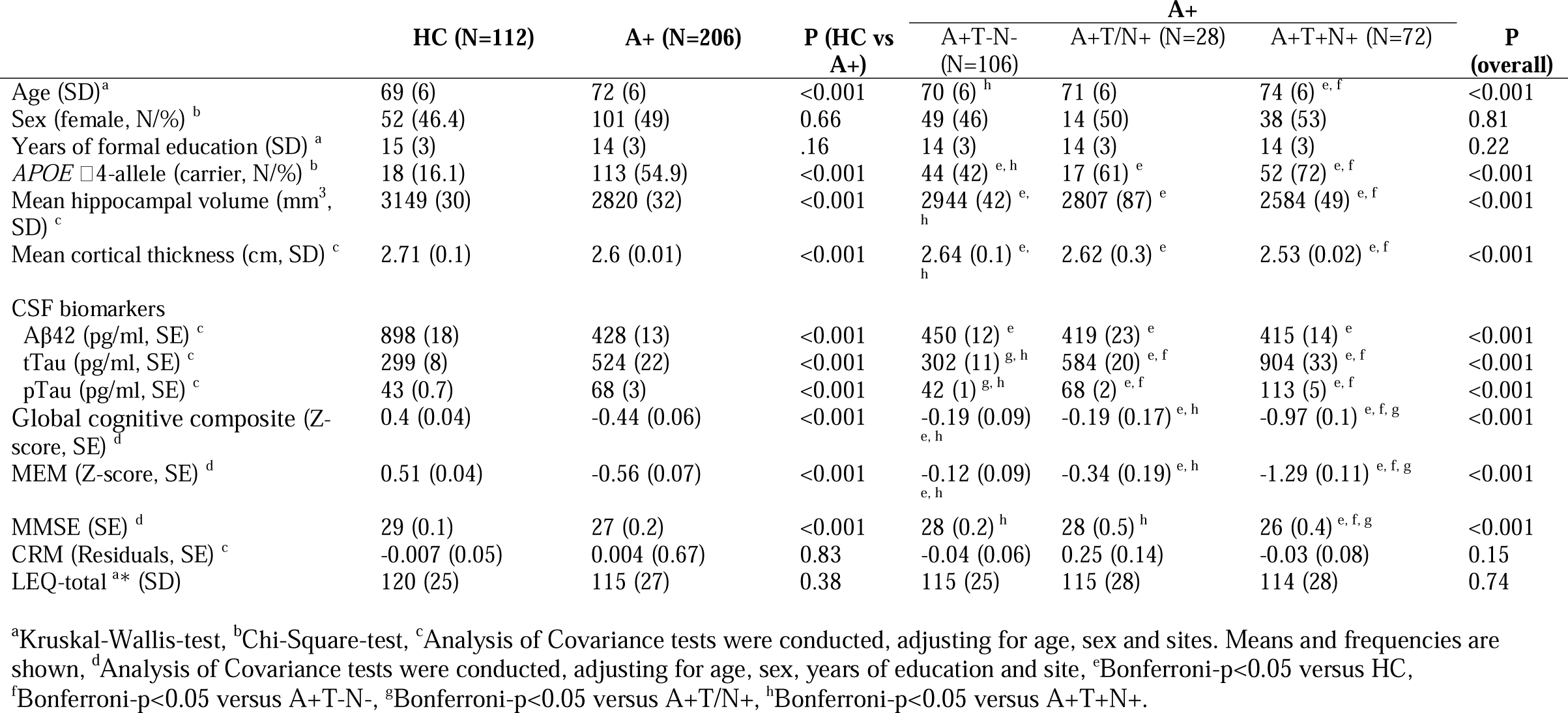
Demographic and clinical characteristics of the study cohort Abbreviations: HC, healthy controls; A+, Amyloid-β positive; Aβ42, Amyloid-beta 42; tTau, total tau; pTau, phosphorylated tau 181; CRM, cognitive reserve marker; MEM, memory cognitive composite score; MMSE, mini-mental state examination; LEQ, lifetime experiences questionnaire; SD, standard deviation; SE, standard error. *Total score of LEQ was available in n=186 (n=63 in HC, n=123 in A+) participants.

### Validation of CRM with educational attainment and lifetime experiences

CRM predicted years of education when analyzing the entire cohort (Fig. S1B, b=0.24, p<0.001, adjusted-R^2^= 0.17) and the HC (b=0.25, p<0.001, adjusted-R^2^= 0.13) and A+ subgroups separately (b=0.25, p=0.01, adjusted-R^2^= 0.2). Furthermore, higher CRM was associated with higher LEQ-total scores in the entire sample (Fig. S1-C, b=0.24, p<0.001, adjusted-R^2^= 0.14) and in the HC (b=0.23, p=0.01, adjusted-R^2^= 0.11) and A+ subgroups (b=0.26, p=0.04, adjusted-R^2^= 0.13).

### Associations of intrinsic network connectivity with MEM

To validate the relationships between INC of RSNs and MEM, we tested the associations between INC of any RSNs and MEM separately. In the entire cohort, a positive association between INC in the DMN and MEM was revealed. Furthermore, higher MEM scores were associated with higher anti-correlation between DMN and SAL (**Table 2 and Fig. 1-A**). In the FPN and the SAL, however, MEM scores were negatively associated with FC in frontal and parietal brain regions (**Table 2 and Fig. 1-B and C**).

**Table 2.**
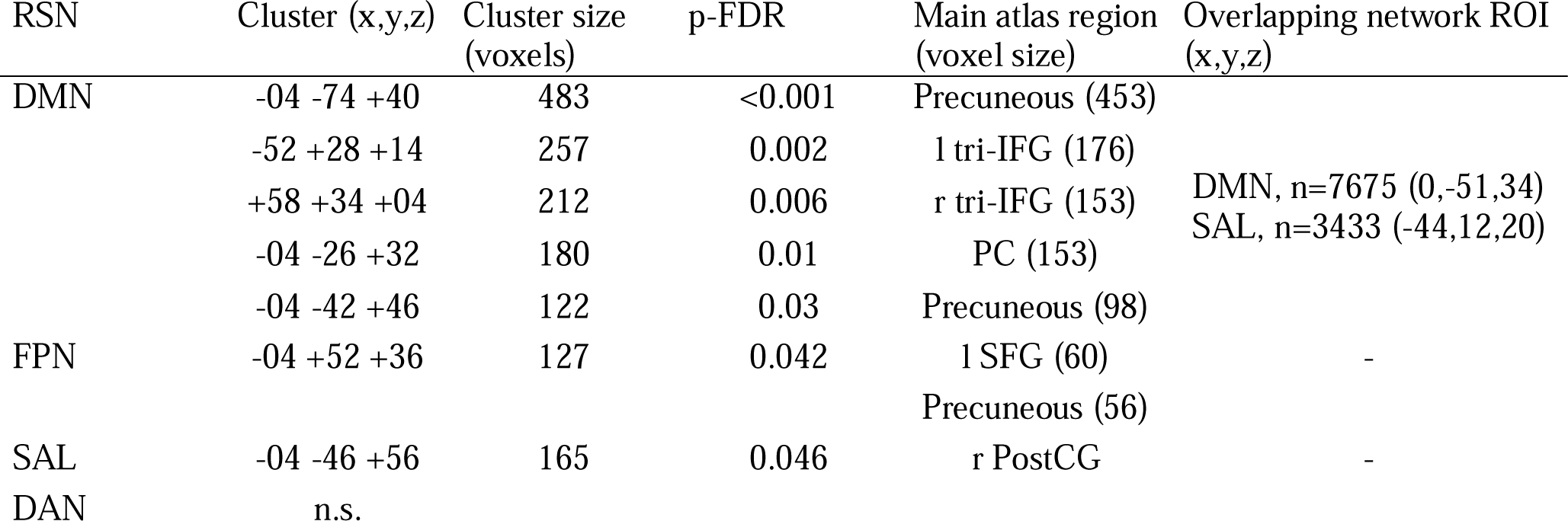
Associations between memory cognitive composite score and intrinsic network connectivity. Results are significant when corrected with gaussian random field theory and with a cluster-level FDR-corrected-p < 0.05. Models were adjusted for age, sex, site and cortical thickness composite score. Anatomic descriptions were made according to the atlas regions in Harvard-Oxford Atlas. Abbreviations: RSN, resting-state network; DMN, default mode network; FPN, frontoparietal network; SAL, salience network, DAN, dorsal attention network; FDR, false-discovery rate; tri-IFG, Inferior Frontal Gyrus, pars triangularis; PC, Cingulate Gyrus, posterior division; SFG, superior frontal gyrus: PostCG, postcentral gyrus; l, left; r, right.

**Figure 1.**
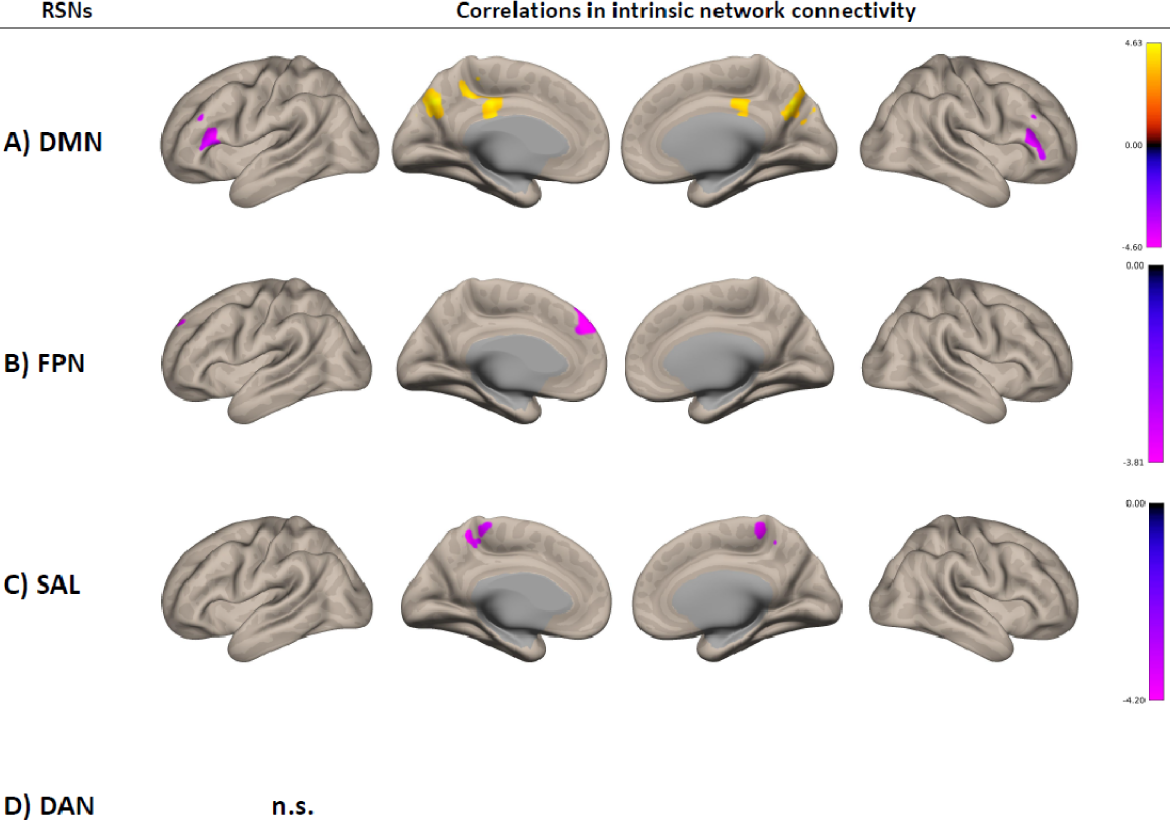
Associations between memory cognitive composite score and intrinsic network connectivity. Results are significant when corrected with gaussian random field theory and with a cluster-level FDR-corrected-p < 0.05. Models were adjusted for age, sex, site and cortical thickness composite score. Anatomic descriptions were made according to the atlas regions in Harvard-Oxford Atlas. The color bar represents T-values. Detailed statistical results are shown in **Table 2**. Abbreviations: DMN, default mode network; FPN, frontoparietal network; SAL, salience network; DAN, dorsal attention network; FDR, false-discovery rate; n.s., not significant.

### Associations between CRM and intrinsic connectivity within cognitive resting-state networks

Higher CRM scores were associated with higher INC within the DMN, particularly in the posterior cingulate cortex and the precuneus (**Table 3 and Fig. 2**-**A**). Additionally, a positive association between the FPN and frontal regions not defined in cognitive RSNs was revealed (**Table 3 and Fig. 2-B**) when tested in the entire cohort. In a subgroup analysis, CRM was associated with a higher anti-correlation between the FPN and DMN in A+T-N- and between the SAL and the DMN in the A+T/N+ group (**Table 3, Fig. 2-B and C**). We found a negative association of INC with CRM in the FPN and in the frontal pole within the A+T/N-group and a negative association between CRM and ICN in the SAL in occipital regions in the A+T/N+ groups, showing no spatial overlap with the cognitive RSNs.

**Table 3.**
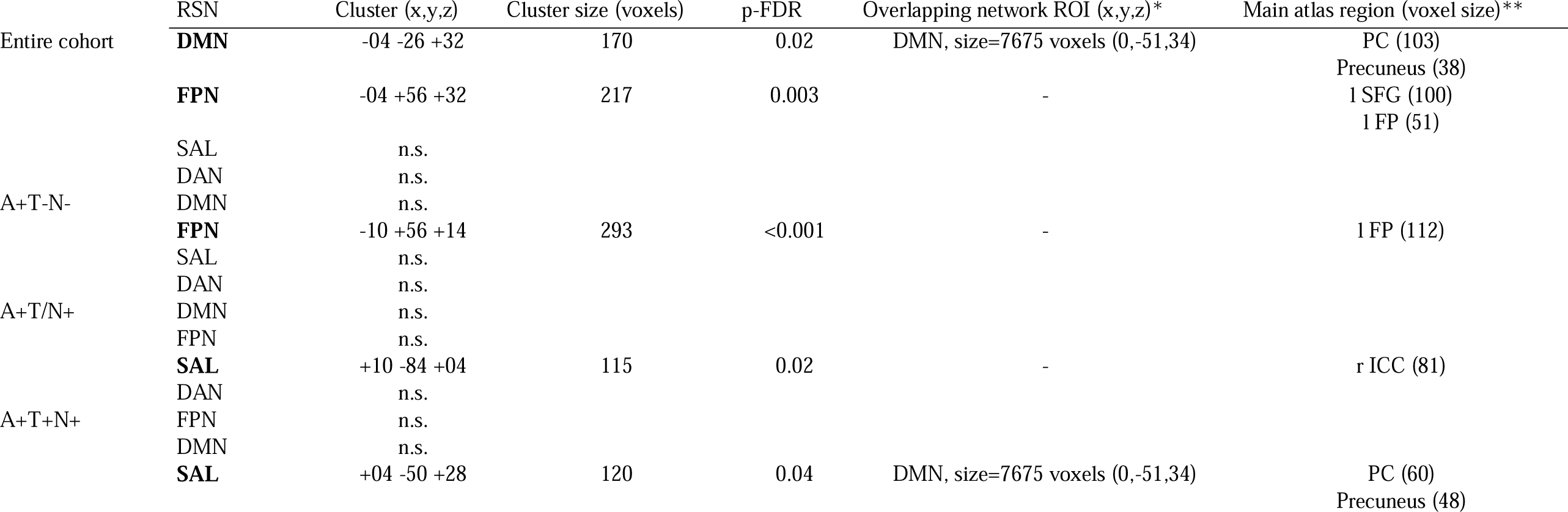
Associations between CRM and intrinsic connectivity of resting-state networks using the general linear models in the entire cohort and in the A/T/N groups. Models were adjusted for age, sex, site, cortical thickness composite score and -in the entire cohort-also for A/T/N group. Results are significant when corrected with gaussian random field theory and with a cluster-level FDR-corrected-p < 0.05. *Network regions from the independent component analysis. **Atlas regions in Harvard-Oxford Atlas. Abbreviations: CRM, cognitive reserve marker; DMN, default mode network; FPN, frontoparietal network; SAL, salience network; ROI, region of interest; FDR, false discovery rate; p-FDR, FDR-corrected p-value; SFG, Superior Frontal Gyrus; FP, Frontal Pole; PC, Cingulate Gyrus, posterior division; ICC, Intracalcarine Cortex; l, left; r, right.

**Figure 2.**
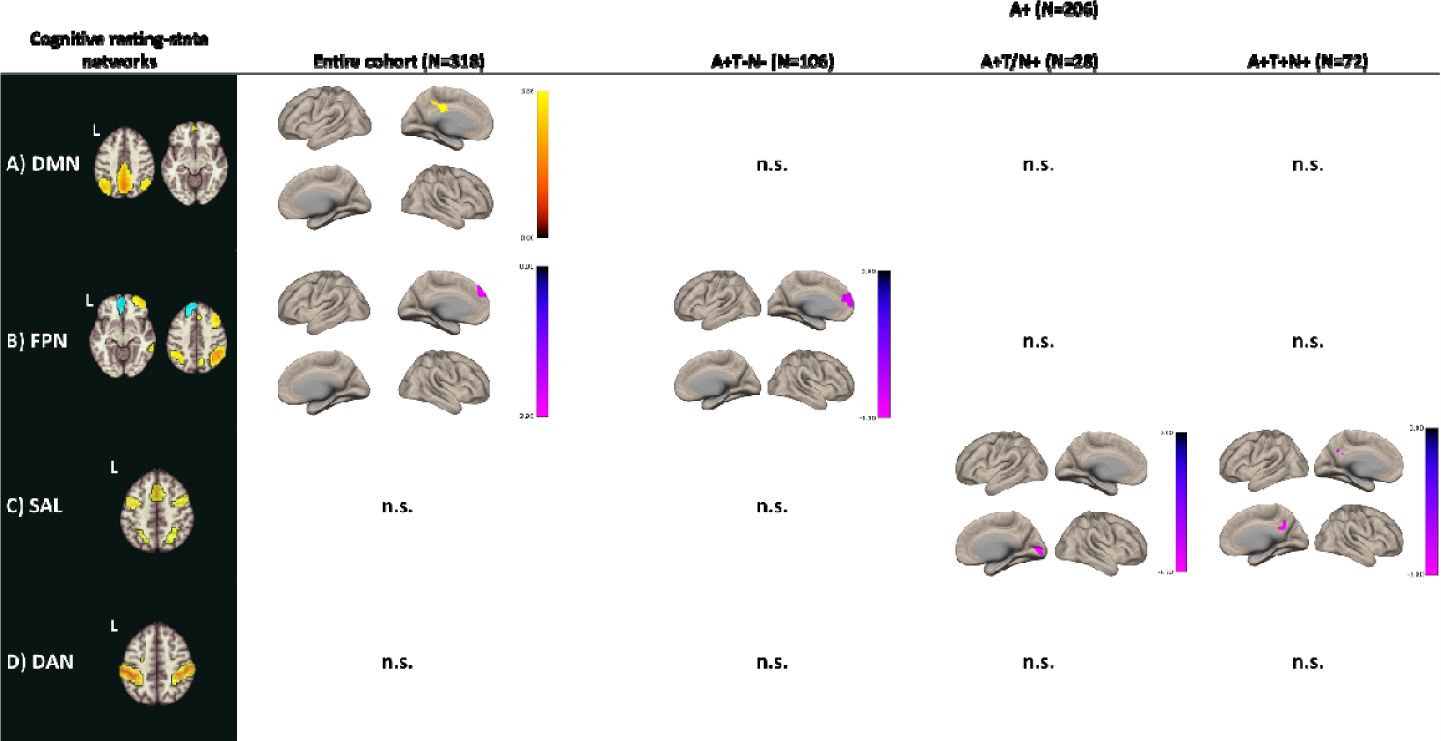
Associations between CRM and intrinsic connectivity of cognitive resting-state networks in the entire cohort and A/T/N subgroups. Models were adjusted for age, sex, site, cortical thickness composite score and in the entire cohort also for A/T/N group. The color bars represent T-values. Results are significant when corrected with gaussian random field theory and with a cluster-level FDR-corrected-p < 0.05. Regions that are identified in RSNs are defined by using the threshold of >2 z-score following the independent component analysis. The results are presented with anatomical descriptions and with exact p-FDR values in **Table 3**. Abbreviations: CRM, cognitive reserve marker; A+, Alzheimer’s disease; DMN, default mode network; FPN, frontoparietal network; SAL, salience network; DAN, dorsal attention network; FDR, false discovery rate; p-FDR, FDR-corrected p-value; n.s., not significant; L, left.

### Associations between CRM and FC of MEM-related seed regions

A seed-to-voxel analysis revealed a negative association of FC between MEM-related regions within the FPN and right parietal regions belonging to the DMN and the FPN (**Table 4, Fig. 3-B**) in the A+ group. Furthermore, the MEM-seed in the FPN showed higher anti-correlation associated with CRM in the precuneus within the DMN in the A+T/N+ group (**Table 4, Fig. 3-B**). We found no associations of CRM with seed connectivity of MEM-related INC regions derived from the DMN, SAL or DAN.

**Table 4.**
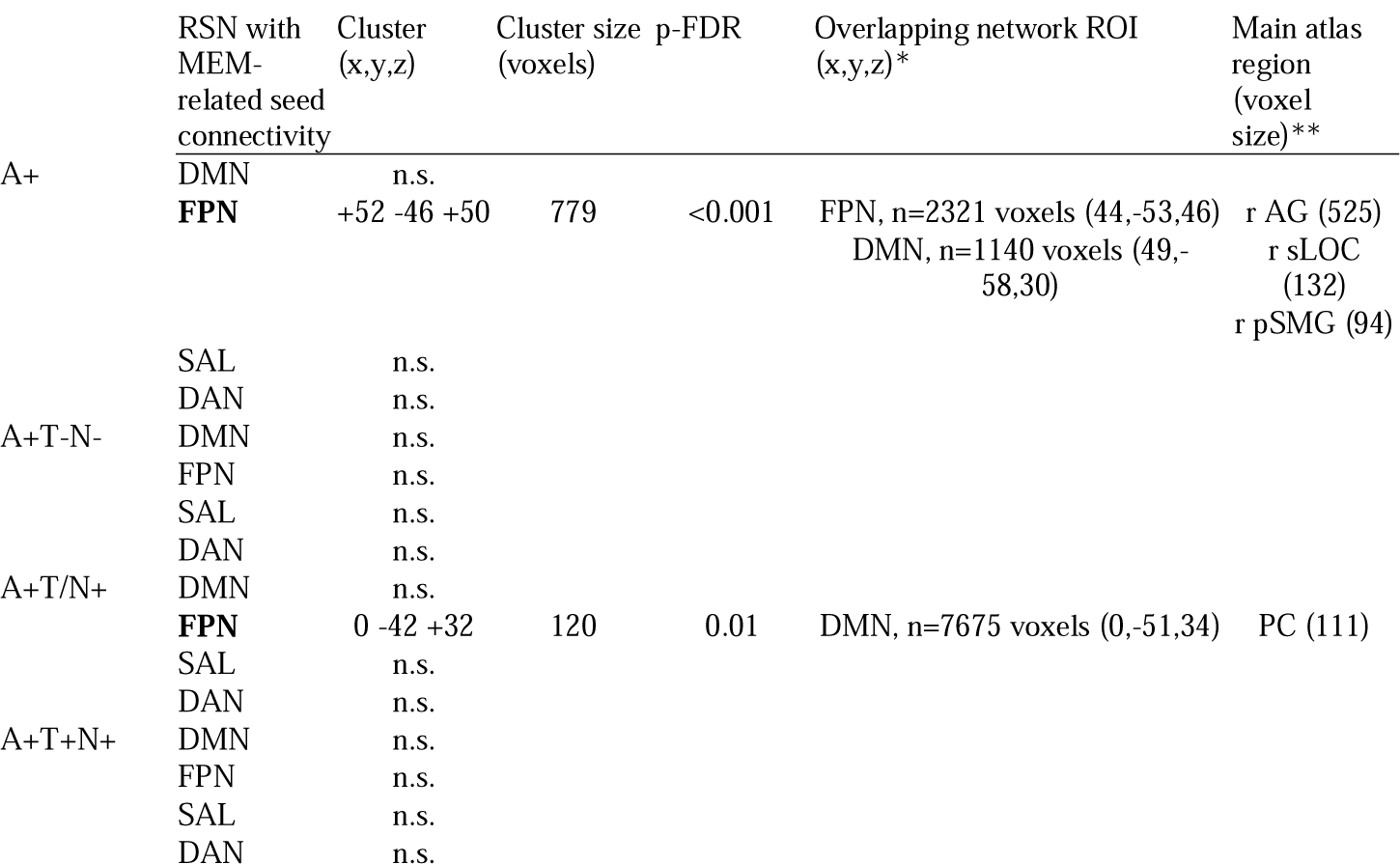
Associations between CRM and memory domain score related connectivity seed for each resting-state network using general linear models in the A+ and in A/T/N groups. Models were adjusted for age, sex, site, cortical thickness composite score and -in the A+-also for A/T/N group. Results are significant when corrected with gaussian random field theory and with a cluster-level FDR-corrected-p < 0.05. *Network regions from the independent component analysis. **Atlas regions in Harvard-Oxford Atlas. Abbreviations: CRM, cognitive reserve marker; A+, Amyloid-β positive; DMN, default mode network; FPN, frontoparietal network; SAL, salience network; ROI, region of interest; FDR, false discovery rate; p-FDR, FDR-corrected p-value;

**Figure 3.**
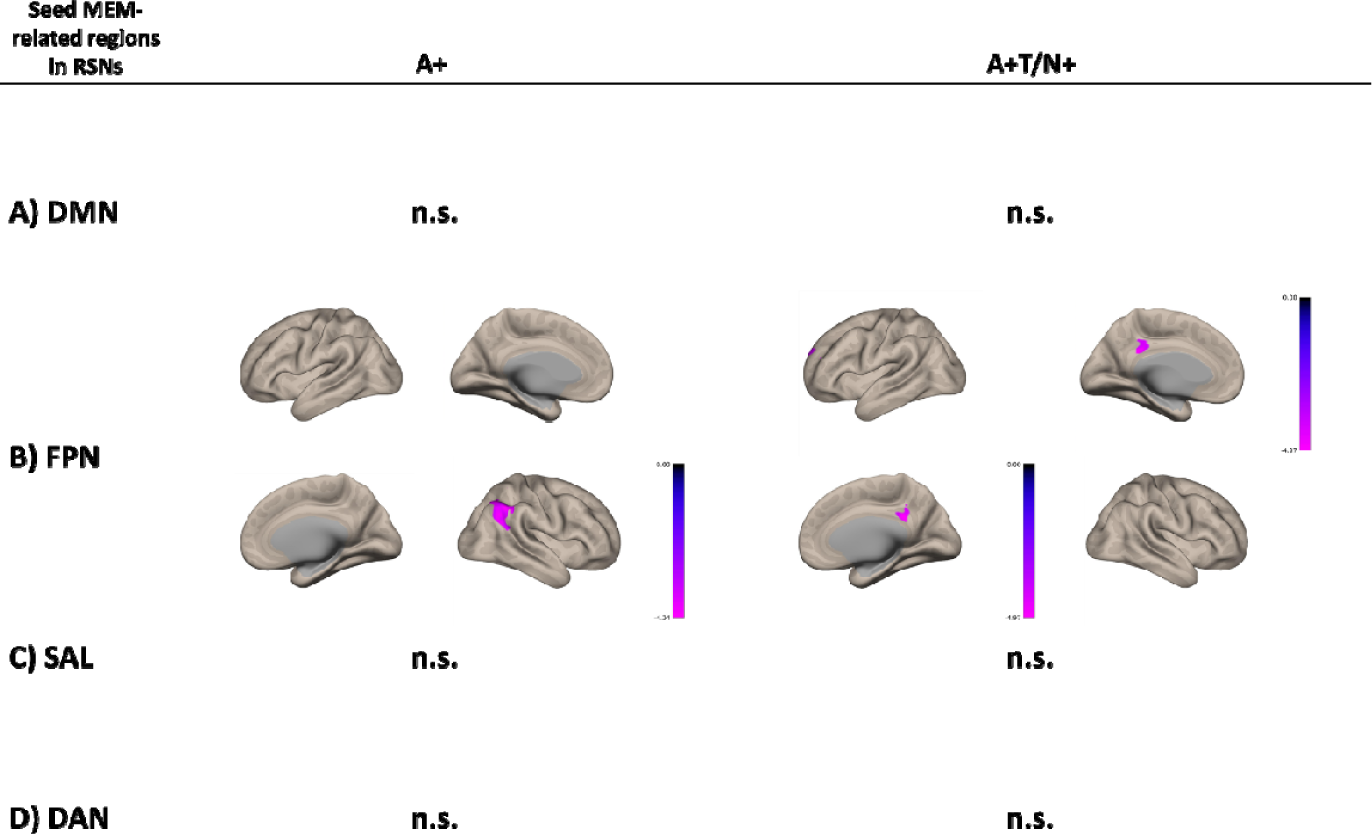
Associations between CRM and functional connectivity of seeds of MEM-related regions in each cognitive resting-state network in the entire cohort and A/T/N subgroups. Models were adjusted for age, sex, site, cortical thickness composite score and in the entire cohort also for A/T/N group. The analyses were conducted separately for each network. The color bar represents T-values. Results are significant when corrected with gaussian random field theory and with a cluster-level FDR-corrected-p < 0.05. The results are presented with anatomical descriptions and with exact p-FDR values in **Table 4**. Abbreviations: CRM, cognitive reserve marker; A+, Amyloid-β positive; DMN, default mode network; FPN, frontoparietal network; SAL, salience network; DAN, dorsal attention network; RSN, resting-state network; n.s., not significant.

## Discussion

The present study provides further evidence on the neural underpinnings of CR estimated using residualized cognitive performance. In Aβ positive individuals higher CRM is associated with INC changes within and between cognitive RSNs, particularly the DMN, the FPN and the SAL. The CRM was associated with commonly used socio-behavioral CR proxies, including years of education, and the LEQ total score, assessing mental activity levels over the lifespan. Our experiment extends previous research by including the aspect of biomarker-defined A/T/N groups, revealing neural correlates of residual CRM, especially in the earlier AD stages (A+T/N- and A+T/N+). This finding supports previous findings suggesting that reserve has its largest impact in the transitional stage between physiological aging and advanced neurodegeneration (2,51).

The observed positive associations between INC changes and memory scores are in line with the literature, showing similar associations in the DMN, particularly in the posterior cingulate cortex and precuneus (44,52). We found an increase in network connectivity in the DMN and FPN with higher performance in the memory domain and higher connectivity between networks of the posterior DMN regions and the FPN. We also found negative associations between memory composite scores and between-network connectivity for SAL-DMN and SAL-FPN and between the FPN and the anterior DMN, while no association was observed between memory and INC in the DAN.

Higher INC in the DMN might contribute to CR when disruptions in the functional network due to AD-related neurodegenerative changes occur. Therefore, the associations between CRM and INC within the DMN may suggest inter-individual variability in network properties, pointing towards a possible neural representation of CR. Previous studies provided evidence on more efficient networks underpinning higher CR, particularly involving regions in the DMN (22,53,54) and FPN (25,55). We observed higher INC in the DMN in subjects with higher CR when tested in the entire cohort. Here, A/T/N groups revealed different associations between INC and CRM, with a higher anti-correlation between DMN and SAL only found in the A+T+N+ group. The INC of the FPN showed a lower FC in the medial frontal region, also part of the DMN (52), suggesting a possible association between CRM and DMN-FPN anti-correlation. In line with this conclusion, a previous study suggested a CR-related higher anti-correlation between DMN and left frontal cortex (i.e, Brodmann area 6/44), a hub region of FPN (24).

Our findings support a major role of the FPN in CR and, more precisely, in neural compensation. In previous work, an association between CR proxies such as education and functional connectivity of FPN were relatively stronger in MCI (51), with no association between the activity of FPN and increased compensation, as no temporary changes in FPN activity were observed with disease progression (25). Similarly, the results of the present study show differences between the entire cohort and A+ individuals, suggesting compensatory changes. A recent interventional study identified the effects of cognitive intervention, showing improved FPN activity and better maintenance of DMN activity in amnestic MCI after a vision-based speed of processing training (56). This pilot study provides an approach to explaining the neural underpinnings of CR and demonstrates the practical value of the concept for developing effective intervention strategies against cognitive decline. Other non-invasive stimulation techniques, such as transcranial magnetic stimulation and focused ultrasound pulse stimulation, may have similar beneficial effects on RSNs (57,58).

A limitation of our work is the cross-sectional study design, precluding firm conclusions on causality. However, due to relatively large group sizes, our results are sufficiently powered to support the validity of the observed associations. Future studies with longitudinal datasets are needed to examine causal relationships between functional network measures and residual CRM. We recommend a further characterization of residual CRM in biomarker-stratified cohorts in future studies. As the residual approach has been studied using different statistical approaches and/or modalities (26), it is less established compared to socio-behavioral proxies of CR; this shortcoming should be addressed in future studies. However, we conducted regression analyses to investigate the associations between CRM and education and lifelong experiences to validate the residual approach.

To conclude, our results advance the understanding of the neurobiological substrates of CR by delineating mechanisms of the neural implementation in functional RSNs. The detailed characterization of CRM-related network differences among individuals with AD pathology and controls will be relevant for the design of future clinical trials and preventive strategies in AD.

## Data Availability

All data produced in the present study are available upon reasonable request to the authors.

## ACKNOWLEDGEMENTS

The DZNE-Longitudinal Cognitive Impairment and Dementia Study (DELCODE) study was funded by the German Center for Neurodegenerative Diseases (Deutsches Zentrum für Neurodegenerative Erkrankungen, DZNE), reference number BN012.

## DISCLOSURES

Ersin Ersoezlue, Boris-Stephan Rauchmann, Maia Tatò, Julia Utecht, Carolin Kurz, Jan Häckert, Selim Guersel, Lena Burow, Gabriele Koller, Sophia Stöcklein, Daniel Keeser, Boris Papazov, Marie Totzke, Tommaso Ballarini, Frederic Brosseron, Katharina Buerger, Peter Dechent, Laura Dobisch, Michael Ewers, Klaus Fliessbach, Wenzel Glanz, John Dylan Haynes, Michael T Heneka, Daniel Janowitz, Ingo Kilimann, Luca Kleineidam, Christoph Laske, Franziska Maier, Matthias H Munk, Oliver Peters, osef PrillerAlfredo Ramirez, Sandra Röske, Nina Roy, Klaus Scheffler, Anja Schneider, Björn H Schott, Annika Spottke, Eike Jakob Spruth, Stefan Teipel, Chantal Unterfeld, Michael Wagner, Xiao Wang, Jens Wiltfang, Steffen Wolfsgruber, Renat Yakupov and Emrah Düzel report no disclosures. Frank Jessen received fees for consultation from Eli Lilly, Novartis, Roche, BioGene, MSD, Piramal, Janssen and Lundbeck. Josef Priller received fees for consultation, lectures and patents from Neurimmune, Axon, Desitin and Epomedics. Robert Perneczky received speaker honoraria and fees for consultation from Janssen, Roche, Biogen, Eli Lilly, Abbott, Schwabe and Grifols.

## Notes

### Clinical Protocols

https://pubmed.ncbi.nlm.nih.gov/29415768/

### Funding Statement

The study was funded by the German Center for Neurodegenerative Diseases (Deutsches Zentrum fuer Neurodegenerative Erkrankungen (DZNE)), reference
number BN012.

### Author Declarations

The study protocol was approved by the ethical committees of the medical faculties of all participating sites: the ethical committees of Berlin (Charité, University Medicine), Bonn, Cologne, Goettingen, Magdeburg, Munich (Ludwig-Maximilians-University), Rostock, and Tuebingen. The process was led and coordinated by the ethical committee of the medical faculty of the University of Bonn. The registration number of the trial at the ethical committee in Bonn is 117/13.

