## Supplemental Figure 1 for "A residual marker of cognitive reserve is associated with resting-state intrinsic functional connectivity along the Alzheimer’s disease continuum"

**Figure S1. A)** Multilinear regression model to estimate cognitive reserve marker in the entire cohort. Multilinear regressions on years of education **(B)** and lifetime experiences questionnaire **(C)** in the entire cohort.

Abbreviations: NPT_global_score, neuropsychological test (global cognitive composite score); edyears, years of education; leq_tot_norm, total lifetime experiences questionnaire score.

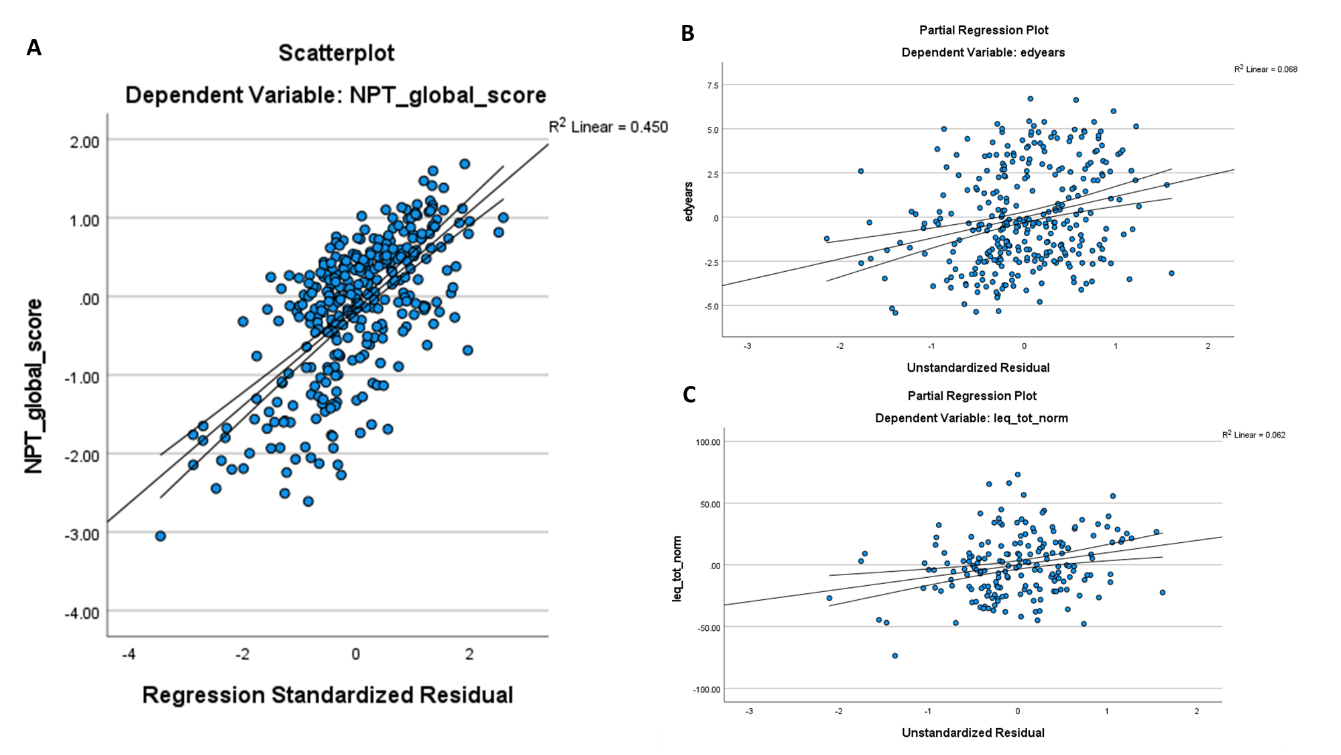
